# Risk factors for household transmission of SARS-Cov-2: a modelling study in the French national population-based EpiCov cohort

**DOI:** 10.1101/2022.10.06.22280739

**Authors:** Sophie Novelli, Lulla Opatowski, Carmelite Manto, Delphine Rahib, Xavier de Lamballerie, Josiane Warszawski, Laurence Meyer, the EpiCov study group

**Author notes:** **Corresponding author:** Sophie Novelli, Inserm U1018, CESP, Hôpital de Bicêtre, 82 rue du Général Leclerc 94276 le Kremlin Bicêtre cedex, France.

## Abstract

**Background:** Households are specific transmission settings, as they involve close and repeated contacts between individuals of different generations. Household surveys provide a unique opportunity to better understand SARS-CoV-2 transmission and the role of individual characteristics.

Here, we assessed the risk of SARS-CoV-2 acquisition from household and community exposure according to age, family ties, and socioeconomic and living conditions using data from the nationwide population-based EpiCov cohort/ORCHESTRA collaboration in November-December 2020.

**Methods:** A history of SARS-CoV-2 infection was defined by a positive Euroimmun Anti-SARS-CoV-2 ELISA IgG result in November-December 2020. We applied stochastic chain binomial models fitted to the final distribution of infections in households to data from 17,983 individuals ≥5 years enrolled from 8,165 households. Models estimated the competing risks of being infected from community and household exposure.

**Results:** Young adults aged 18-24 years had the highest risk of extra-household infection (8.9%, [95% credible interval, Crl]: 7.5 – 10.4), whereas the oldest (>75) and the youngest (6-10) had the lowest risk, 2.6% (1.8 – 3.5) and 3.4% (1.9 – 5.2), respectively. Extra-household infection was also independently associated with socioeconomic conditions. Within households, the probability of person-to-person transmission increased with age: 10.6% (5.0 – 17.9) among 6-10-year-olds to 43.1% (32.6 – 53.2) among 65-74-year-olds. It was higher between partners 29.9% (25.6 - 34.3) and from mother to child 29.1% (21.4 – 37.3) than between individuals related by other family ties.

**Conclusion:** In 2020 in France, the main factors identified for extra-household infection were age and socioeconomic conditions. Intra-household infection mainly depended on age and family ties.

**Key Messages:**

- Young adults aged 18-24 years had the highest probability of extra-household SARS-Cov-2 acquisition over the year 2020: 8.9%, 95% credible interval (95%Crl) 7.5 – 10.4.
- The probability of extra-household infection increased with family income and population density in the municipality of residence and was higher in the French regions most affected by the waves of SARS-CoV-2.
- When estimating the probability of person-to-person transmission of SARS-CoV-2, the 65-74 year-olds had the highest susceptibility, i.e. the highest probability of SARS-CoV-2 acquisition when exposed to an infected household member (22.1%, 16.4 – 28.2)
- The probability of transmission was the highest between partners (29.9%, 25.6 – 34.3). The probability of transmission was higher from mother to child than from father to child: 29.1%, (21.4 – 37.3) and 14.0% (5.9 – 22.8), respectively. The probability of transmission from child to parent was higher from children <12 years than for older children: 11.8% (2.5 – 25.1) and 4.1% (0.9 – 9.0), respectively.

## INTRODUCTION

Monitoring households, where individuals of different generations have close and repeated contacts can help us understand the transmission of pathogens. Regarding COVID-19, household studies have provided accumulating evidence that susceptibility to SARS-CoV-2 infection is lower for children (<8-10 years of age), relative to adults, and higher among those >60-65 years relative to younger/middle-aged adults ^1,2^. However, evidence is still lacking regarding risk for newborns, young children, and adolescents, while these age groups are characterized by highly heterogeneous behavior and social contact patterns.

Based on a population-based serosurvey in Geneva, Switzerland over April-June 2020, Bi et al. identified a reduced risk of within-household infection among young children (5-8 years of age) and young people (10-19) relative to middle-aged adults (20-49)^3^. This study was however carried out a time of particularly low prevalence among children across Europe ^4–8^ after a long period of school closure during which children had limited interactions outside of the home. limiting the possibility of studying routes of infection and infectivity among children. From June-December 2020, data from a Canadian pediatric cohort highlighted higher infectivity among children than adolescents ^9^. Importantly despite growing evidence of social disparities in the risk of SARS-CoV-2 infection ^10–14^ socio-economic factors were rarely evaluated at the household level.

The EpiCov cohort is a rich nationwide population-based serosurvey in France, with a household survey performed in November-December 2020, when schools were open and vaccination had not yet been implemented. Serological assays are used to measure antibody responses in blood samples as the sign of a previous SARS-Cov-2 infection, while virological tests detect ongoing infection. While these later may miss mild or asymptomatic infections ^15^, serological tests remain sensitive over a longer period ^16^ and thus consist in complementary tools to define infection history and study SARS-CoV-2 transmission. Here, we used mathematical modelling to assess, from the Epicov data, the effects of age, family ties, and living and socio-economic conditions on the risk of SARS-CoV-2 acquisition from both household and community exposure.

## METHODS

### The EpiCov cohort

EpiCov is a national random population-based cohort that combines serological testing and longitudinal follow-up. It aims to analyze both the impact of living conditions on the dynamics of the epidemic and the impact of the epidemic on health and living conditions in France. In May 2020, 371,000 individuals aged ≥15 years living in mainland France or three of the five French overseas territories were randomly selected from the Fidéli administrative sampling frame. This database is considered to be quasi-exhaustive for the population living in France ^17^. The survey design was defined to ensure overrepresentation of the less densely populated departments and lower income households, for which lower response rates were expected. Selected individuals were contacted to undergo a web/telephone questionnaire. The survey design, multimodal data collection have been detailed elsewhere ^11^.

### Household study design

In November 2020, a 20% subsample of EpiCov participants was randomly drawn to be part of a household study. Eligible participants were offered home capillary blood self-sampling for SARS-CoV-2 serology for all household members, i.e., any person living at the same address aged >6 years. Only one household member per household, i.e., the one initially sampled to be part of the EpiCov cohort, completed the questionnaire. He/she was defined as the respondent member. Households for whom the respondent was aged <17 years or living in a household of >9 members were not proposed to participate in the household sub-study.

### Epidemiological context of the household survey

This household survey aimed to capture infections that had occurred from the start of the pandemic in France (February-March 2020) to November-December 2020. This period covers the first two waves of the COVID-19 pandemic, mostly caused by the wildtype virus before the alpha variant gradually became dominant after its introduction at the end of 2020 ^18^ and before the start of the vaccination campaign on December 27, 2020 ^19^. The epidemiological evolution during the year 2020 in France has been described elsewhere ^12^ and is briefly detailed in Supplementary Note 1.

### Laboratory analyses

Dried-blood spots were collected on 903Whatman paper (DBS) kits sent to each participant who agreed to blood sampling and mailed to one of the three participating biobanks (Bordeaux, Amiens, Montpellier) to be punched using a PantheraTM machine (Perkin Elmer). Eluates were processed in the virology laboratory (Unité des Virus Emergents, Marseille) with a commercial ELISA kit (Euroimmun^®^, Lübeck, Germany) for the detection of anti-SARS-CoV-2 antibodies (IgG) against the S1 domain of the viral spike protein (ELISA-S), according to the manufacturer’s instructions.

### Outcome

SARS-Cov-2 seropositivity was defined as an ELISA-S IgG ratiol1≥1.1, according to the ratio threshold supplied by the manufacturer, and considered as the main criteria.

### Exposures

Collected data included the number, age, and gender of all individuals living in the household and the decile income of the household per capita. We also considered the administrative region, the population density in the municipality of residence, whether the housing was overcrowded, defined as housing with <18 m^2^ per inhabitant, and whether the neighborhood was defined as socially deprived according to national definitions for prioritizing targeted socio-economic interventions.

### Ethics

The survey was approved by the CNIL (the French data protection authority) (MLD/MFI/AR205138) and the local ethics committee (Comité de Protection des Personnes Sud Mediteranée III 2020-A01191-38). The survey was also reviewed by the “Comité du Label de la Statistique Publique”. The serological results were sent to the participants by post with information on how to interpret the individual test results.

### Statistical analysis

We analyzed all households for which serostatus was available for all members. Because serostatus of young children was not available in EpiCov, households with children ≤5 years of age were removed. We applied stochastic chain binomial transmission models to the final distribution of infections in households ^3,20^. The models accounted for competing risks between community and household infection by considering all possible sequences of SARS-Cov2 introduction and subsequent transmission events within a household. We estimated: 1) the probability that a susceptible individual *i* was infected from extra-household exposure since the beginning of the pandemic to the time of the serosurvey and 2) the probability that a susceptible individual *i* was infected from a single infectious household member *j*; note this was not the overall probability of being infected within the household but a probability of person-to-person transmission of SARS-CoV-2. For the purposes of simplicity, we considered the serological status to be a perfect marker of having been infected and neglected the occurrence of false positives or false negatives. We also neglected the possibility that individuals could have been infected several times.

We investigated factors associated with the risk of extra- and within-household SARS-Cov-2 acquisition. They included the characteristics of the susceptible individual *i* and the potential infector *j*, as well as the living and socioeconomic conditions of the household (see Supplementary Note 2). The variables considered for modeling the probability of extra-household acquisition were: age and gender of the susceptible member, family income, population density in the municipality of residence, and living in a socially deprived neighborhood. We also considered immigration status, this variable being collected for the respondent. We accounted for spatial heterogeneity in the extra-household probability, adjusting for the administrative region. For the probability that an infectious household member *j* infects a susceptible household member *i*, we considered the following covariates: age and gender of the susceptible individual *i*, age and gender of the potential infector *j*, family ties between *i* and *j* (i.e., whether *j* was *i*’s partner, parent, child, sibling, grandparent, grandchild, or other), household size, accommodation type, and number of rooms. We also tested whether demographic and socioeconomic factors (immigration status, family income, population density in the municipality of residence, and living in a socially deprived neighborhood) and administrative region modulated the risk of within-household transmission.

We built a series of models including various combinations of these variables. Model parameters were estimated by Bayesian inference via Markov Chain Monte Carlo (MCMC) sampling. Model fits were compared using the widely applicable information criterion (WAIC) ^21^. Given the very low percentage of missing data for the considered variables (<4%), we run complete case analysis.

We adapted the code written by Bi et al. ^3^ and available in open access to the EPICOV data. Models were implemented in the Stan probabilistic programming language using the rstan R package (version 2.21.2). We used weakly informative priors on all parameters to be normally distributed on the logit scale, with a mean of 0 and a standard error of 1.5. We ran four chains of 1,500 iterations each with 500 warm-up iterations. We assessed convergence visually and using the Gelman-Rubin Convergence Statistic (R-hat). We report estimates as the medians of the posterior samples with their 95% credible interval (Crl), i.e., the 2.5th and 97.5th percentiles of their distribution.

## RESULTS

### Study population

Among the 22,118 households in mainland France drawn from the EpiCov cohort to be part of the household sub-study in November 2020, we analyzed 17,983 individuals belonging to 8,165 households for which the serostatus of all members was available (Figure 1). The median date of blood sampling was November 27^th^, 2020 (interquartile range, IQR: 23th November–6th December). The median age of the participants was 52 years (IQR, 28–64), and male-female sex-ratio 0.93. The household characteristics are shown in Table 1. Most of the households were one-person (25.9%) or two-person (47.4%), which differed from the general population (36.9% and 32.6% one and two-person households in 2019, respectively ^22^). The main family structures were couples without (43.2%) and with children (23.8%). Young people (6-24 years) and middle-aged adults (35-54) mainly lived in households with >2 members, whereas 25-34 and >55-year-olds mainly lived in two-person households (Table S1). The proportion of one-person households was the highest among 25-34-year-olds (21.1%) and >75-year-olds (21.0%).

**Table 1.**
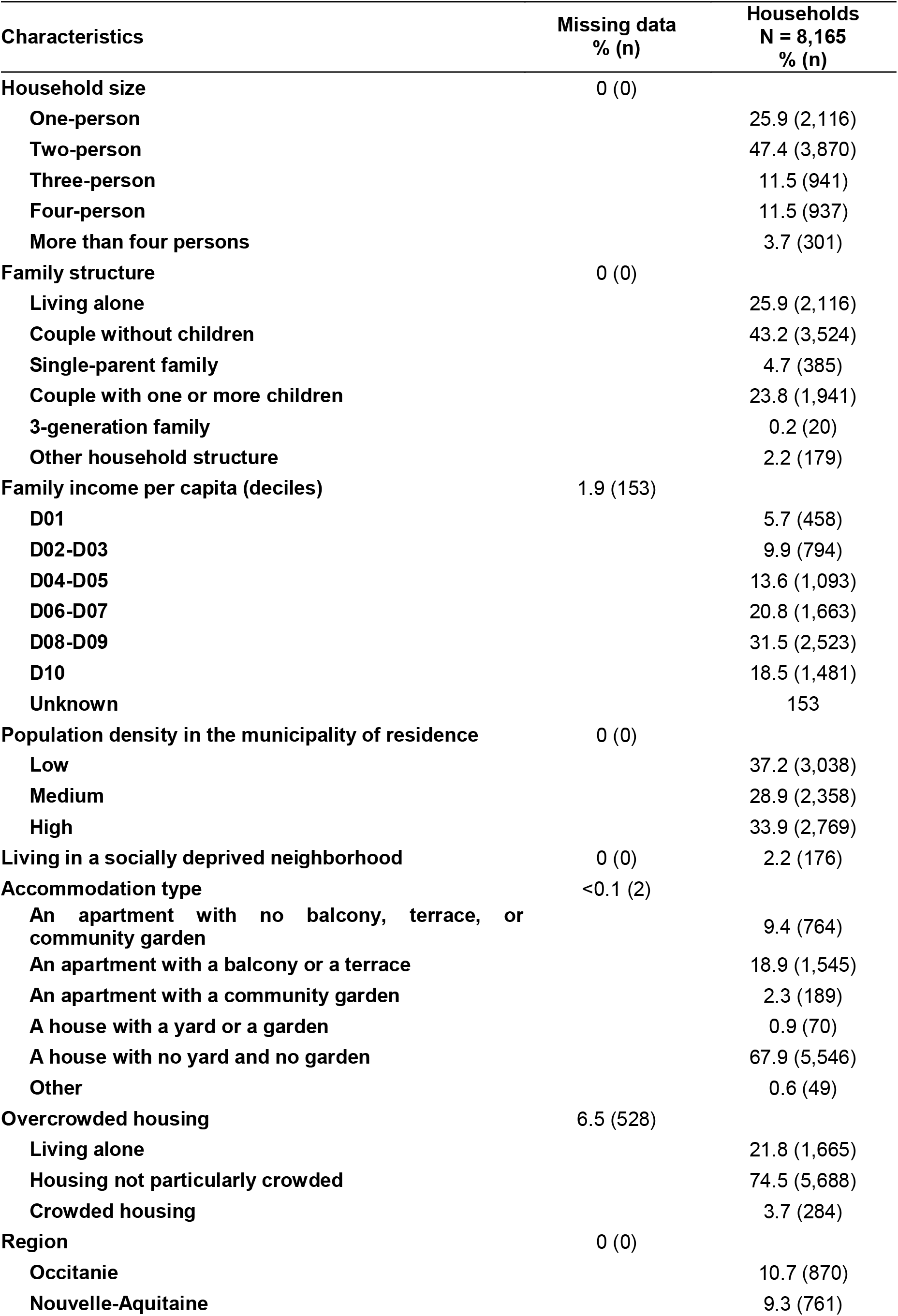
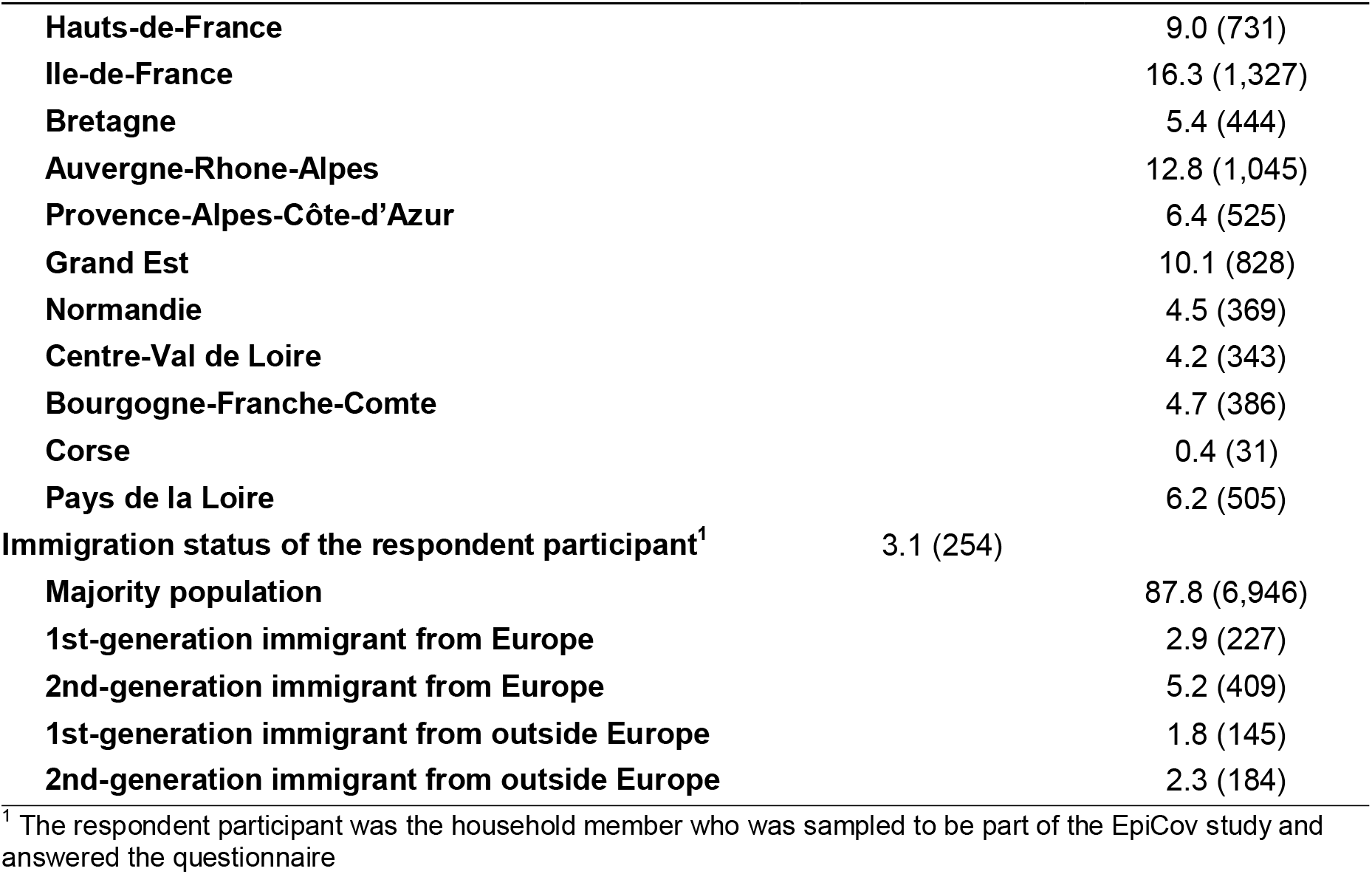
Household characteristics.

**Figure 1.**
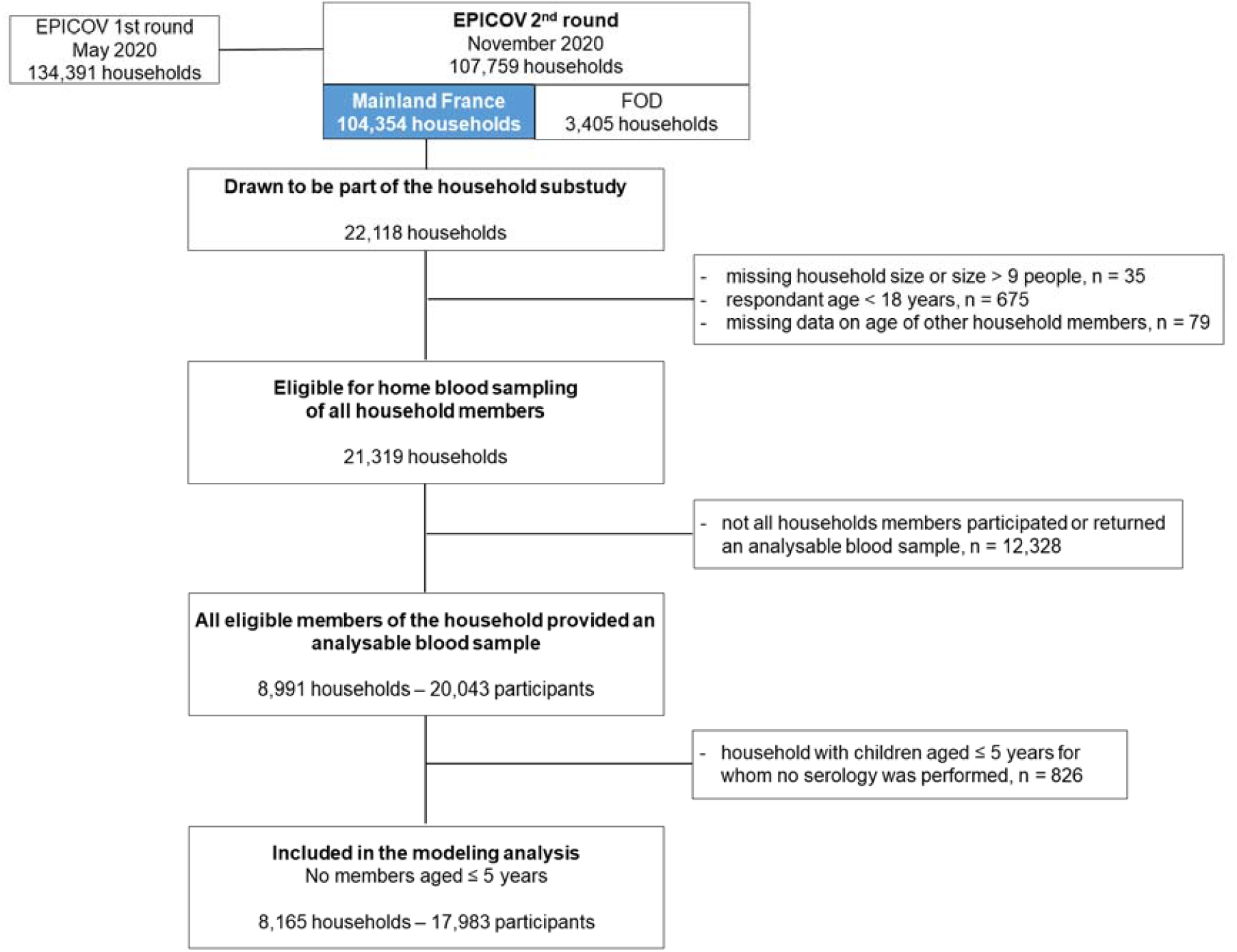
Flowchart of enrolled participants. Abbreviations: FOD, French Overseas Departments

### Seroprevalence and household final attack rates

At the individual level, overall seroprevalence did not differ by gender but varied with age, increasing from 5.5% among 6-10-year-olds to 7.5% among 11-14-year-olds and 7.7% among 15-17-year-olds, before peaking at 10.7% among 18-24-year-olds and then decreasing to 3.0% among those aged ≥75 (Figure 2A,B).

**Figure 2.**
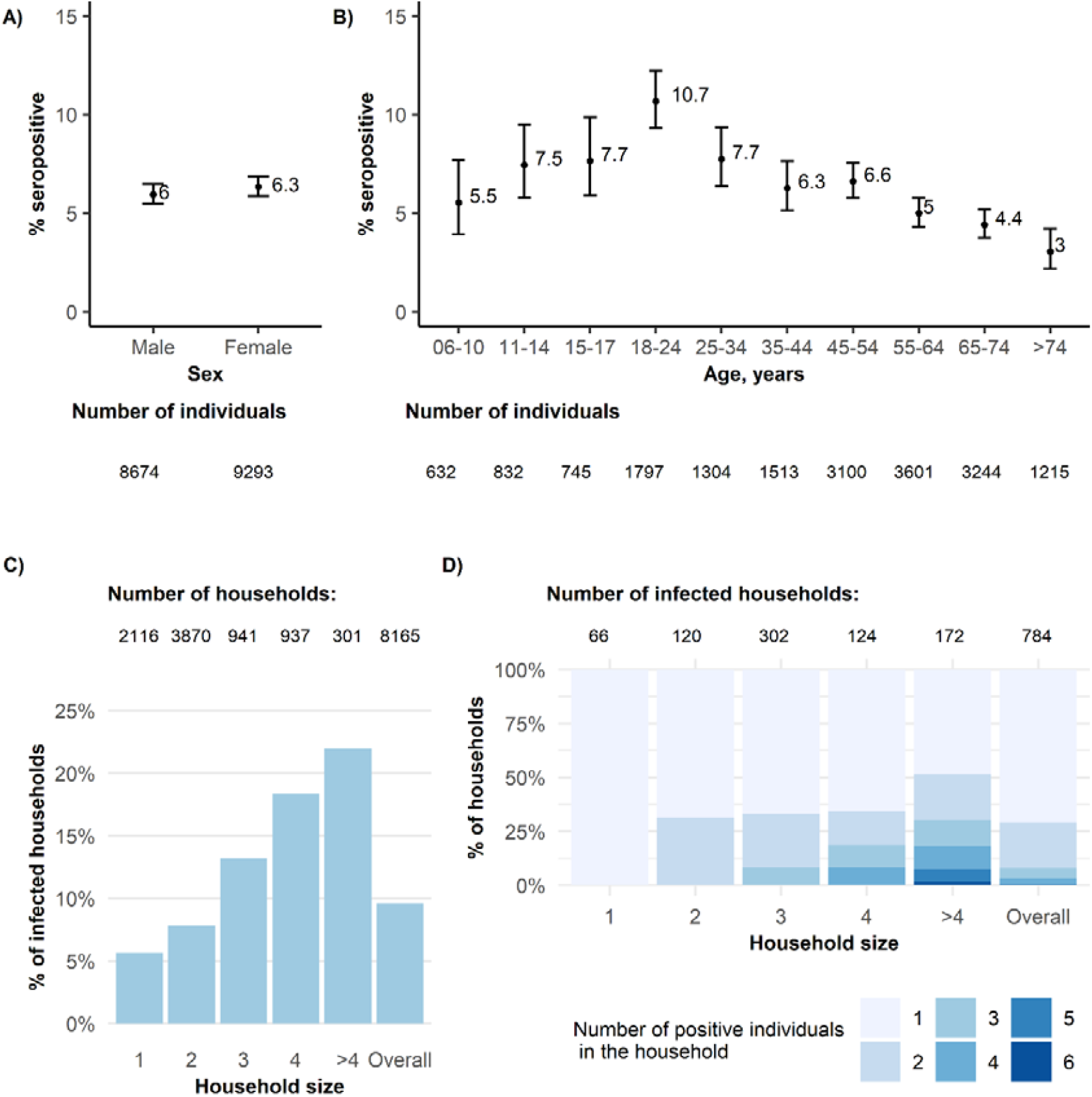
Study population: (A) Proportion of infected households, i.e., with at least one positive case, according to household size. (B) Among households with a least one positive individual, total number of positive individuals in the household according to the size of the household. Example: Among households with three members with at least one positive individual, 67% had only one positive household member, 25% two positive members, and 8% three positive members. (C) and (D) Seroprevalence (95% CI) according to age and sex.

The proportion of households with at least one seropositive member was 9.6% overall and increased from 5.1% for one-person households to 23.1% for households with ≥5 members (Figure 2C, Table S2). Most infected households counted only one or two positive individuals, even in households >3 members (Figure 2D). Among households with at least one positive individual, the proportion of positive members was 52% overall and decreased with household size, from 66% in two-person households to 47% in three-person households, to 40% in 4 and >4-person households.

### Factors associated with the acquisition of SARS-CoV-2

We estimated the overall cumulative probability of infection from extra-household exposure from the start of the pandemic in France through the time of the survey to be a median of 4.5% (95% CrI 4.2–4.9). This probability varied with age: young adults aged 18-24 years had the highest probability of extra-household infection and those aged ≥75 had the lowest (Figure 3A).

**Figure 3.**
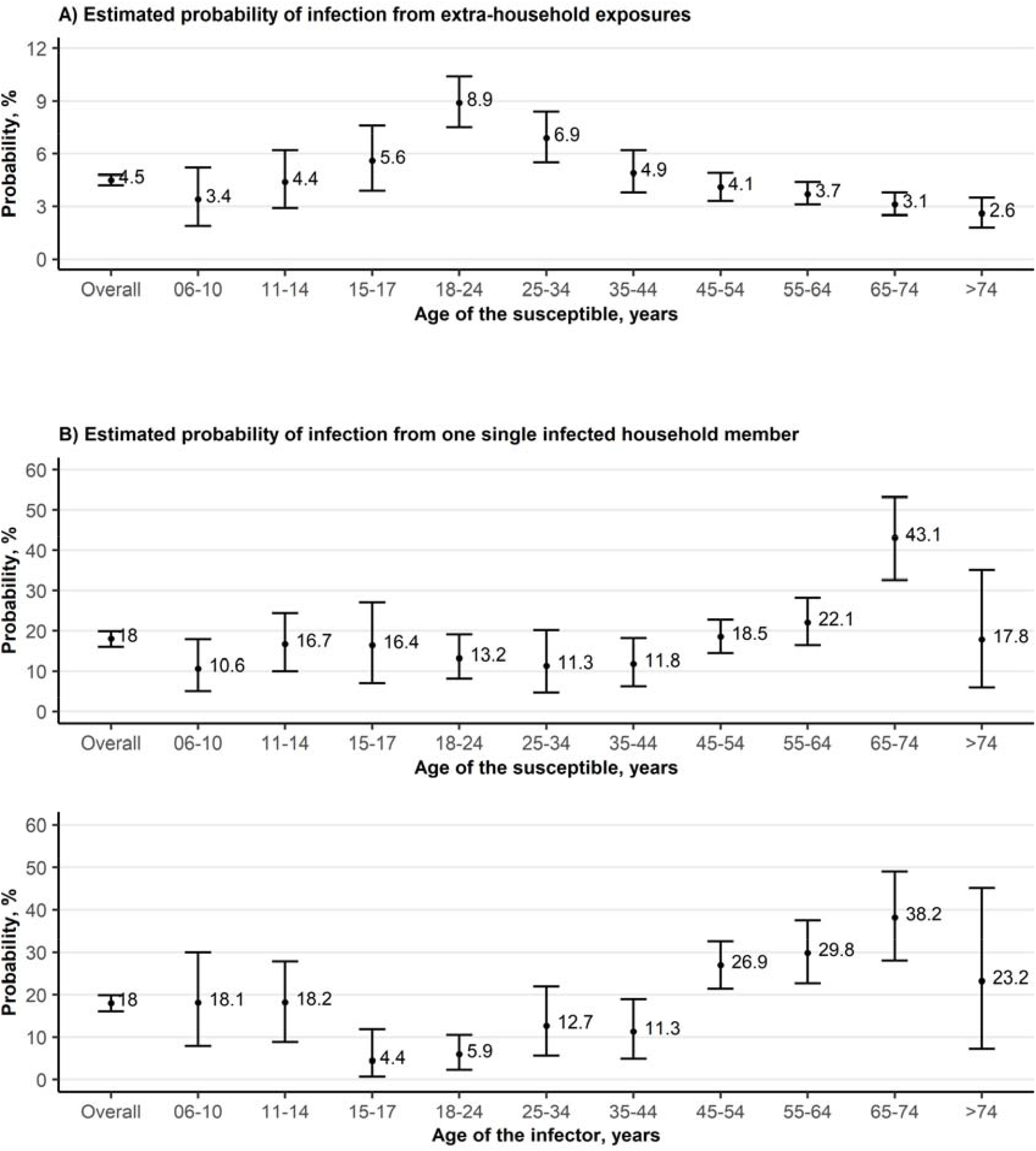
Estimated probabilities (%) of extra and intra-household infection by age. Median, 95% credible interval

The estimated median probability of person-to-person transmission between household members was 17.9% (16.0–19.9) overall. It varied with age of the susceptible: susceptibility was the highest among 65-74-year-olds, and the lowest among 6-10-year-old (Figure 3B). Models allowing for differential risk of transmission by the age of the infector showed the lowest infectivity for the 15-17 and 18-24 age groups (Figure 3C). The probability of person-to-person transmission between household members was 29.5% (24.3–34.9) in two-person households, and decreased to 15% in larger households. These overall estimates concealed heterogeneous probabilities of transmission according to family ties. The probability of transmission was the highest between partners (29.9%, 25.6–34.3), which was consistent with the estimate in two-person households (Figure 4A,B). The probability of transmission was higher from mother to child (29.1%, 21.4–37.3) than from father to child (14.0%, 5.9– 22.8). From child to parent it was of 11.8% (2.5–25.1) for children <12 years of age and decreased to 4.1% (0.9–9.0) for children ≥12 years of age. The limited number of three-generation families in the sample led to wide credible intervals when estimating the transmission risk between grandchildren and grandparents. Family income, population density in the municipality of residence, immigration status, and region were not associated with the risk of person-to-person transmission between household members (Table S3).

**Figure 4.**
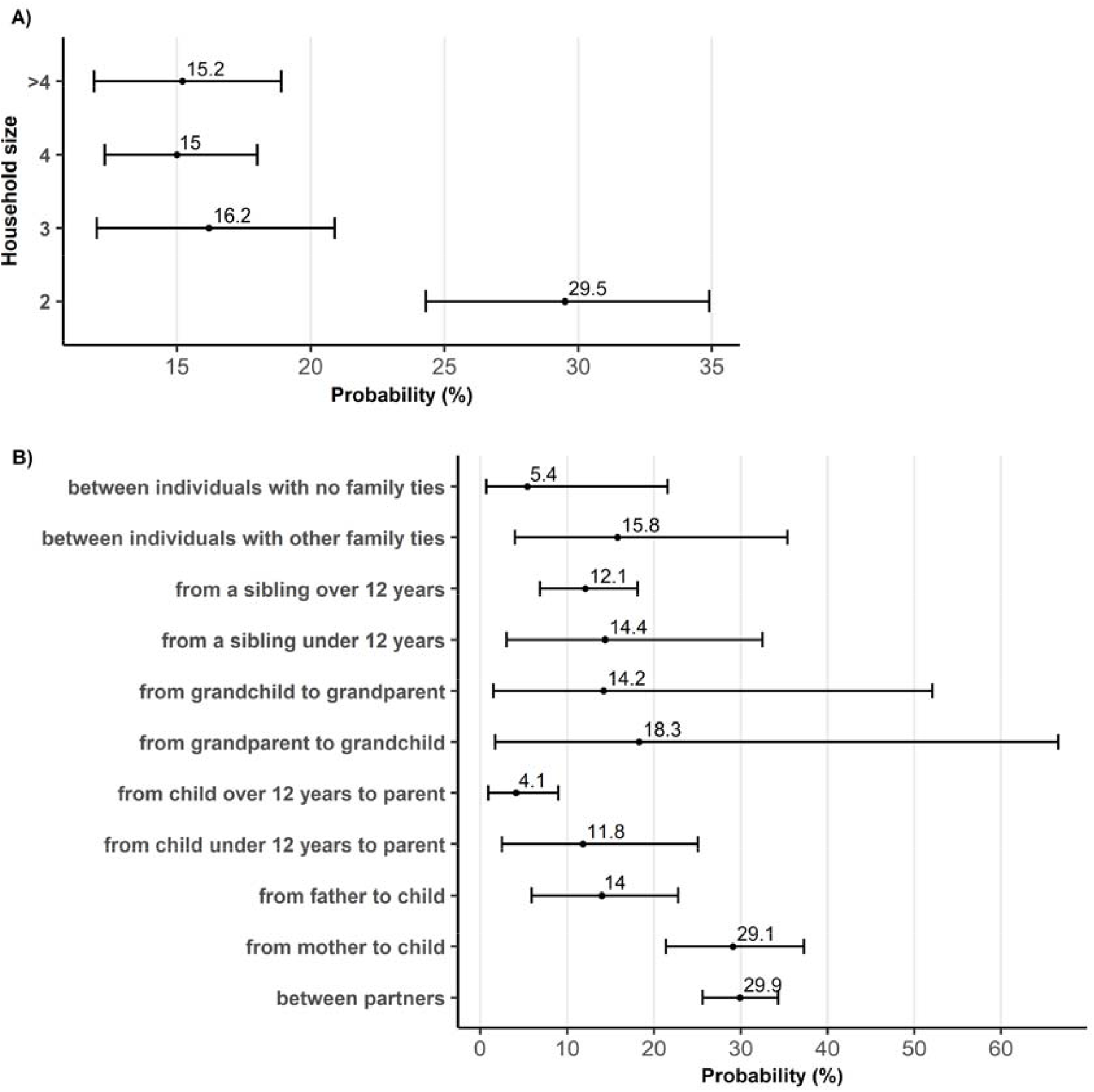
Estimated probabilities (%) of person-to-person transmission according to (A) household size and (B) family ties. Median, 95% credible interval

In multivariate analysis, the probability of being infected from extra-household exposure was adjusted for the age of the susceptible individual, family income, population density in the municipality of residence, immigration status of the respondent, and the administrative region of residence. The association with age remained similar: young people (18-24-year-olds) and middle-aged adults (25-34 and 35-44-year-olds) had a higher risk of extra-household SARS-CoV-2 acquisition than that of 55-64-year-olds (Table 2). The probability of extra-household infection increased with family income (highest for the two highest deciles compared to the central deciles) and population density in the municipality of residence. Individuals in households in which the respondent participant was a first-or second-generation immigrant from outside Europe had a higher risk of extra-household SARS-CoV-2 acquisition in univariate analysis. This effect was tempered after adjusting for socioeconomic factors. Finally, as expected, the regions with the highest SARS-CoV-2 incidence at the time of the survey were those with the highest probability of extra-household infection.

**Table 2.** Factors associated with a risk of extra-household and intra-household infection.

| Variables | Infection from extra-household exposures |  | Infection from one single infected household member |  |
| --- | --- | --- | --- | --- |
|  | Univariate analysis | Multivariate analysis | Univariate analysis | Multivariate analysis 1 |
|  | OR 95%CrI | OR 95%CrI | OR 95%CrI | OR 95%CrI |
| **Age of the susceptible, years** |  |  |  |  |
| 06-10 | 0.9 (0.5 - 1.5) | 1.0 (0.5 - 1.6) | 0.4 (0.2 - 0.8) | 0.4 (0.1 - 1.2) |
| 11-14 | 1.2 (0.8 - 1.8) | 1.1 (0.6 - 1.6) | 0.7 (0.4 - 1.3) | 0.9 (0.3 - 2.1) |
| 15-17 | 1.5 (1.0 - 2.2) | 1.4 (0.9 - 2.1) | 0.7 (0.3 - 1.4) | 1.1 (0.4 - 2.8) |
| 18-24 | 2.5 (2.0 - 3.3) | 2.4 (1.8 - 3.1) | 0.5 (0.3 - 0.9) | 0.9 (0.4 - 2.0) |
| 25-34 | 1.9 (1.4 - 2.5) | 1.6 (1.2 - 2.2) | 0.5 (0.2 - 1.0) | 0.5 (0.2 - 1.1) |
| 35-44 | 1.3 (1.0 - 1.8) | 1.4 (1.0 - 1.9) | 0.5 (0.2 - 0.9) | 0.5 (0.2 - 1.1) |
| 45-54 | 1.1 (0.8 - 1.4) | 1.2 (0.9 - 1.6) | 0.8 (0.5 - 1.3) | 1.0 (0.6 - 1.9) |
| 55-64 | ref | ref | ref | ref |
| 65-74 | 0.8 (0.6 - 1.1) | 0.9 (0.7 - 1.2) | 2.7 (1.5 - 4.7) | 1.9 (1.0 - 3.5) |
| >74 | 0.7 (0.5 - 1.0) | 0.7 (0.5 - 1.0) | 0.8 (0.2 - 2.0) | 0.7 (0.2 - 2.1) |
| **Sex of the susceptible** |  |  |  |  |
| Female | ref |  | ref |  |
| Male | 0.9 (0.8 - 1.1) |  | 0.7 (0.5 - 1.0) |  |
| **Age of the infector, years** |  |  |  |  |
| 06-10 |  |  | 0.5 (0.2 - 1.1) |  |
| 11-14 |  |  | 0.5 (0.2 - 1.1) |  |
| 15-17 |  |  | 0.11 (0.02 - 0.34) |  |
| 18-24 |  |  | 0.15 (0.05 - 0.31) |  |
| 25-34 |  |  | 0.3 (0.1 - 0.7) |  |
| 35-44 |  |  | 0.3 (0.1 - 0.6) |  |
| 45-54 |  |  | 0.9 (0.5 - 1.4) |  |
| 55-64 |  |  | ref |  |
| 65-74 |  |  | 1.5 (0.8 - 2.6) |  |
| >74 |  |  | 0.7 (0.2 - 2.0) |  |
| **Sex of the infector** |  |  |  |  |
| Female |  |  | ref |  |
| Male |  |  | 0.8 (0.5 - 1.3) |  |
| **Family ties** |  |  |  |  |
| Between partners |  |  | ref | ref |
| From mother to child |  |  | 1.0 (0.6 - 1.5) | 1.2 (0.5 - 2.8) |
| From father to child |  |  | 0.4 (0.1 - 0.7) | 0.34 (0.06 - 1.04) |
| From child <12 years old to parent |  |  | 0.31 (0.06 - 0.81) | 0.63 (0.07 - 2.16) |
| From child ≥12 years old to parent |  |  | 0.10 (0.02 - 0.24) | 0.27 (0.09 - 0.57) |
| From grandparent to grandchild |  |  | 0.53 (0.04 - 4.78) | 0.63 (0.04 - 6.67) |
| From grandchild to grandparent |  |  | 0.39 (0.04 - 2.59) | 0.50 (0.04 - 4.50) |
| From a sibling <12 years old |  |  | 0.39 (0.07 - 1.14) | 0.8 (0.1 - 3.0) |
| From a sibling ≥12 years old |  |  | 0.3 (0.2 - 0.6) | 0.4 (0.1 - 1.0) |
| Between individuals with other family ties |  |  | 0.4 (0.1 - 1.3) | 0.5 (0.1 - 1.5) |
| Between individuals with no family ties |  |  | 0.13 (0.02 - 0.66) | 0.18 (0.02 - 1.03) |
| **Immigration status** |  |  |  |  |
| Majority population | ref | ref | ref |  |
| 1st-generation immigrant from Europe | 0.8 (0.5 - 1.2) | 0.8 (0.5 - 1.2) | 1.0 (0.4 - 2.1) |  |
| 1st-generation immigrant from outside Europe | 1.6 (1.0 - 2.4) | 1.3 (0.8 - 2.0) | 0.6 (0.2 - 1.2) |  |
|  | Univariate analysis | Multivariate analysis | Univariate analysis | Multivariate analysis 1 |
|  | OR 95%CrI | OR 95%CrI | OR 95%CrI | OR 95%CrI |
| 2nd-generation immigrant from Europe | 1.0 (0.7 - 1.4) | 1.0 (0.7 - 1.4) | 1.3 (0.7 - 2.3) |  |
| 2nd-generation immigrant from outside Europe | 1.6 (1.0 - 2.3) | 1.4 (0.9 - 2.0) | 1.3 (0.7 - 2.4) |  |
| **Family income** |  |  |  |  |
| D01 (lowest) | 1.2 (0.8 - 1.8) | 1.0 (0.7 - 1.5) | 0.5 (0.2 - 1.2) |  |
| D02-D03 | 1.0 (0.7 - 1.4) | 0.9 (0.7 - 1.3) | 1.3 (0.7 - 2.3) |  |
| D04-D05 | ref | ref | ref |  |
| D06-D07 | 1.2 (0.9 - 1.5) | 1.2 (0.9 - 1.6) | 1.1 (0.7 - 1.9) |  |
| D08-D09 | 1.3 (1.1 - 1.7) | 1.3 (1.0 - 1.7) | 1.1 (0.7 - 1.7) |  |
| D10 (highest) | 1.7 (1.3 - 2.2) | 1.6 (1.2 - 2.1) | 1.4 (0.9 - 2.3) |  |
| **Population density in the municipality of residence** |  |  |  |  |
| Low | ref | ref | ref |  |
| Medium | 1.2 (1.0 - 1.4) | 1.1 (0.9 - 1.3) | 1.0 (0.7 - 1.4) |  |
| High | 1.6 (1.4 - 1.9) | 1.2 (1.0 - 1.4) | 1.0 (0.7 - 1.4) |  |
| **Household size** |  |  |  |  |
| 2 |  |  | ref | ref |
| 3 |  |  | 0.5 (0.3 - 0.7) | 0.8 (0.4 - 1.4) |
| 4 |  |  | 0.4 (0.3 - 0.6) | 0.8 (0.4 - 1.3) |
| >4 |  |  | 0.4 (0.3 - 0.6) | 0.9 (0.5 - 1.7) |
| **Accommodation type** |  |  |  |  |
| A house with a yard or a garden |  |  | ref |  |
| A house with no yard or garden |  |  | 1.5 (0.4 - 4.0) |  |
| An apartment with a balcony or a terrace |  |  | 1.3 (0.9 - 1.8) |  |
| An apartment with a community garden |  |  | 0.8 (0.1 - 2.5) |  |
| An apartment with no balcony, terrace or community garden |  |  | 1.1 (0.5 - 2.1) |  |
| Other |  |  | 0.9 (0.2 - 3.1) |  |
| **Overcrowded housing** |  |  |  |  |
| Housing not particularly crowded |  |  | ref |  |
| Crowded housing |  |  | 1.2 (0.8 - 1.7) |  |
| **Region** |  |  |  |  |
| Auvergne-Rhone-Alpes | 1.9 (1.3 - 2.9) | 1.8 (1.2 - 2.7) |  |  |
| Bourgogne-Franche-Comte | 1.1 (0.7 - 1.9) | 1.1 (0.7 - 1.9) |  |  |
| Bretagne | 0.6 (0.3 - 1.0) | 0.5 (0.3 - 0.9) |  |  |
| Centre-Val de Loire | ref | ref |  |  |
| Corse | 0.24 (0.02 - 1.22) | 0.28 (0.03 - 1.62) |  |  |
| Grand Est | 1.5 (1.0 - 2.3) | 1.5 (1.0 - 2.3) |  |  |
| Hauts-de-France | 1.8 (1.2 - 2.7) | 1.8 (1.2 - 2.8) |  |  |
| Ile-de-France | 2.3 (1.6 - 3.5) | 1.8 (1.2 - 2.6) |  |  |
| Normandie | 1.1 (0.7 - 1.9) | 1.1 (0.6 - 1.8) |  |  |
| Nouvelle-Aquitaine | 0.8 (0.5 - 1.3) | 0.8 (0.5 - 1.3) |  |  |
| Occitanie | 0.9 (0.6 - 1.4) | 0.9 (0.6 - 1.4) |  |  |
| Pays de la Loire | 1.0 (0.6 - 1.6) | 1.0 (0.6 - 1.6) |  |  |
| Provence-Alpes-Cote d'Azur | 0.8 (0.5 - 1.4) | 0.8 (0.5 - 1.3) |  |  |

Abbreviations: OR odds ratio; 95% CrI, 95% credibility interval; ref, reference

Concerning intra-household transmission, the probability of person-to-person transmission was adjusted for the age of the susceptible and his/her family link with the potential infector. Those aged 65-74 years had twice the odds of being infected from a single infected household member than 55-64-year-olds (aOR 1.9, 95%CrI 1.0–3.5), whereas 6-10-year-olds appeared to have a lower risk of infection (aOR 0.4, 0.1–1.2), but the 95% credible interval was wide. The risk of transmission from mother to child was similar to that between partners aOR, 1.2 (0.5–2.8), whereas it was lower from father to child than from mother to child (aOR 0.34, 0.06–1.04). The risk of transmission from child to parent decreased with increasing age of the child: aOR, 0.63 (95%CrI, 0.07–2.16) for children <12 years and 0.27 (0.09–0.57) for children ≥12 years, using transmission between partners as the reference. When adjusting for family ties, household size was no longer associated with the risk of person-to-person transmission.

Interestingly, living condition variables, i.e., the type of accommodation and overcrowded housing, were not associated with the risk of person-to-person transmission in univariate analysis. The introduction of these two variables into the multivariate analysis did not change the odds ratio estimates for age or family ties.

### Proportion of intra-household infections

Using the posterior distribution of parameters from the final multivariate model, we simulated the predicted sources of infection for all individuals in the study (see Supplementary Note 1). We estimated that 25.5% (95%CrI, 25.1–25.8) of all infections were caused by another household member. This proportion increased with household size, from 22.2% (21.6–22.7) for two-person households to 44.0% (42.5–45.5) for households with ≥5 members (Figure 5).

**Figure 5.**
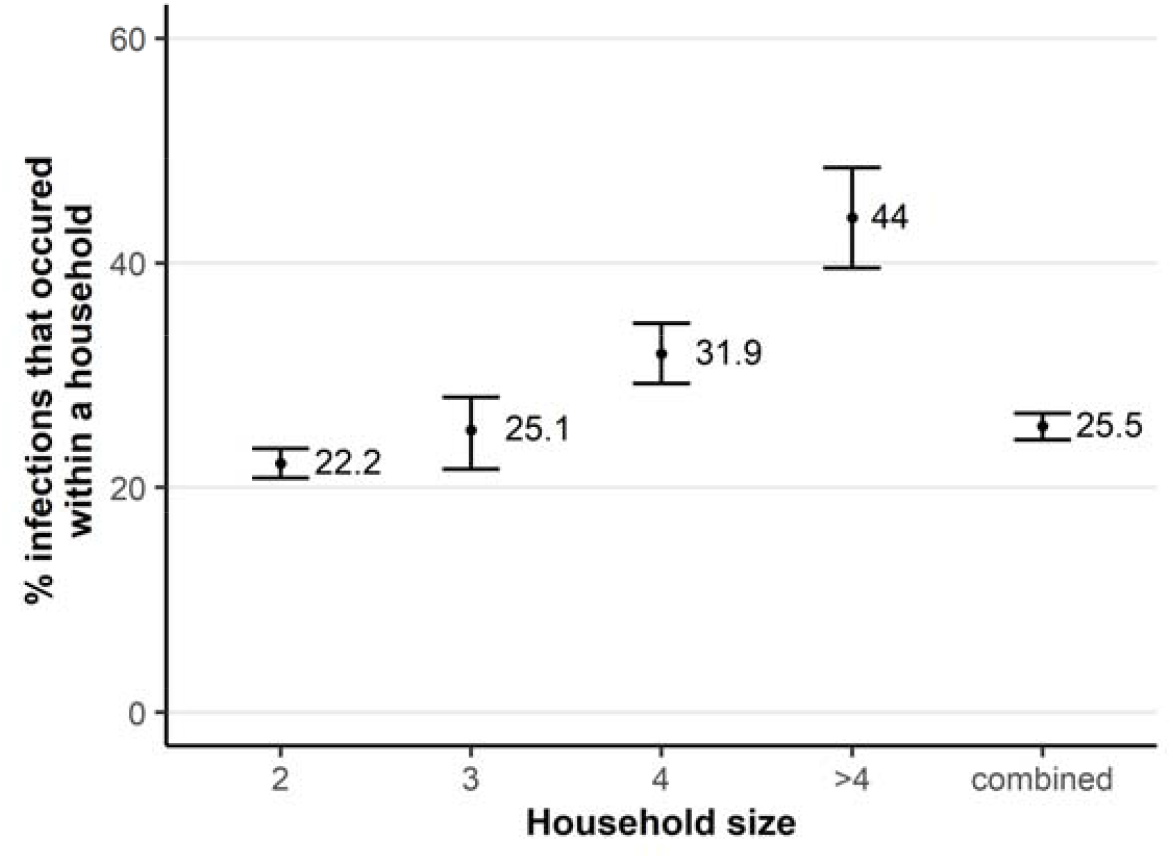
Estimated proportion of infections that occurred within a household depending on household size. Median, 95% credible interval

## DISCUSSION

Using serological data from the French nationwide population-based EpiCov cohort, we estimated the risk of SARS-CoV-2 acquisition from the community and the person-to-person transmission risk within households in 2020. Participants of all ages >5 years were included and we particularly investigated the effect of family composition, socioeconomic factors, and living conditions on within-household SARS-CoV-2 transmission. Based on simulations, we found that household transmission represented one quarter of SARS-CoV-2 infections. The main factors for extra-household infection were age and demographic and socioeconomic factors (i.e., family income, population density in the area of residence, and region), whereas intra-household infection mainly depended on the age of individuals and family ties between them.

Our results regarding an age pattern of SARS-Cov-2 transmission was consistent with previous household studies^3,23^. Young adults had the highest probability of extra-household infection, probably reflecting a higher intensity of social interaction in this age group. The highest probability of being infected when exposed to an infectious individual at home was obtained for the 65-74 year-olds, which may be explained by either a higher susceptibility to virus infection or by more time spent at home relative to the younger age groups. Surprisingly, participants >74 did not have a particularly high risk of being infected by an infectious family member at home. This may reflect a potentially higher level of preventive measures in these populations. A higher level of waning antibodies in the oldest individuals may have also led to an underestimation of the infection rates in this age group ^24^. Importantly, individuals leaving in nursing homes were not included in the EpiCov study. Age-specific probabilities of person-to-person transmission result from a combination of biological effects (immune response and viral shedding) and behavioral factors (differences in social exposure and the frequency and intensity of contacts between age groups in the population). In models accounting for family ties, the probability of within-household transmission was the highest between partners and from mother to child. Lower values were obtained from child to parents. This is consistent with the higher secondary infection rates for spouses than for other adult relationships, which were reported previously ^25,26^. We found a higher probability of transmission to parents from the <12 year-old children than from older children, probably reflecting heterogeneity in contact patterns between individuals within families. Overall, our results suggest that, in a context where schools were open, adults rather than children were more likely to be infected outside and introduced the virus to the household, leading to transmission to children.

Family income and population density in the municipality of residence were associated with the probability of extra-but not intra-household infection, which allows a better understanding of the association of these factors with the seroprevalence previously described in the EpiCov cohort ^11,12^. Surprisingly, we did not find an association between within-household person-to-person transmission and overcrowded housing or accommodation type, by opposition to some previous studies which reported associations between housing surface areas and secondary infection rates in households ^27,28^. This difference may be due to the fact that relatively few dwellings were overcrowded in our sample. Isolation measures taken in households after a member became infected may also explain our results but this information was not available.

The probability of person-to-person transmission was inversely related to household size. It was 30% in two-person households, which corresponds to the estimated probability of transmission between partners. It decreased to 15-16% in larger households, which is an average of all the probabilities of transmission between household members that ranged from 4 to 30% according to families ties. When adjusting for family ties, household size was no longer associated with the risk of person-to-person transmission.

Interestingly, from our simulations we estimated that household transmission represented one quarter of SARS-CoV-2 infections in 2020. This value depended on the person-to-person probability of transmission within households, as well as the structure of the population: the larger the household size and the number of positive household members, the more likely sequences involving one or more within-household infection events. In France, near 50% of households are two-persons. Our study population mainly consisted of one- and two-person households 73%. The contribution of household transmission would be expected to be far higher in settings were larger households are more frequent, as the estimated risk of household transmission increased from 22.2% in two-person households to 44.0% in households with ≥5 members.

Among the main strengths of our study, in addition to its nationwide dimension, is that it covers a period of interest when schools were no longer closed in France, starting from autumn 2020, and children and young people were therefore more exposed to SARS-CoV-2 in the community. In addition, we accounted for competing risks between extra- and intra-household exposure for SARS-CoV-2 acquisition and the possibility of tertiary or longer transmission chains within households by using chain binomial models.

Some limitations need to be mentioned. First, despite the size of our study, one of the largest in Europe, its statistical power was limited for the analysis of specific interactions, e.g. between grandparents and grandchildren. Second, antibody responses are generally sustained over the first four months following infection but may wane afterwards. The study taking place in fall 2020, some infections that occurred early in the first wave of the pandemic may have been missed ^24^. Third, the possibility of re-infection was neglected here. Given that the same viral strain circulated at this time of the pandemic, this phenomenon was considered infrequent ^24^. Finally, lack of availability of serological status in children <5 years in the EpiCov cohort impeded us from investigating the role of very young children in SARS-CoV-2 transmission. More studies should be carried out in this specific group in the future.

In conclusion, our study provides estimates of the risk of acquisition of SARS-CoV-2 within and outside the household over the first two waves in France in 2020. The probability of person-to-person transmission within households was estimated to be 18% overall, and varied highly depending on the age of the individuals and the family ties. Since November 2020, the epidemiological context has drastically changed, with both the emergence of variants of concern with increased transmissibility and widespread implementation of vaccination. Our study brings new insights into the understanding of factors associated with the heterogeneity of SARS-CoV-2 transmission. It also illustrates and highlights the strength of population-based serosurveys to assess the relative contribution of household and community transmission, which can be extended to other respiratory viruses.

## Data Availability

Anonymous aggregated data for the first round are available online. The EpiCov dataset is available for research purpose concerning the first round, and will be available by March 2022 concerning the second round for research purpose on CASD (https://www.casd.eu/) after submission to approval of French Ethics and Regulatory Committee procedure (Comite du Secret Statistique, CESREES and CNIL). Access to anonymized individual data underlying the findings may be available before the planned period, on request to the corresponding author, to be submitted to approval of ethics and reglementary Committee for researchers who meet the criteria for access to data.

## Authors contribution

JW and LM conceived and designed the epidemiological study. CM, XL and DR contributed to data collection. SN and LO conceived the modelling analysis. SN performed the analysis, supervised by LO. SN, LM and LO interpreted the results. SN wrote the first draft of the paper. All authors wrote, read and approved the final manuscript.

## For the EpiCov study group

Josiane Warszawski, Nathalie Bajos (Co-Principal investigator), Guillaume Bagein, François Beck, Emilie Counil, Florence Jusot, Nathalie Lydié, Claude Martin, Laurence Meyer, Philippe Raynaud, Alexandra Rouquette, Ariane Pailhé, Delphine Rahib, Patrick Sillard, Alexis Spire.

## Acknowledgments

The views expressed in this paper are the sole responsibility of the author and the Commission is not responsible for any use that may be made of the information it contains. We sincerely thank all the participants in the EpiCoV study.

We warmly thank the INSERM staff, including, in particular, Carmen Calandra, Karim Ammour, Jean-Marc Boivent, Jean-Marie Gagliolo, Frédérique Le Saulnier, and Frédéric Robergeau, who worked with considerable dedication and commitment to make it possible to develop, in record time, and to maintain all regulatory, budgetary, technical, and logistical aspects of the EpiCov study.

We warmly thank the staff of Santé publique France, and especially Lucie Duchesne, who played a major role in organisation and quality assurance for the seroprevalence component of the EpiCov study.

We thank the CRB biobanks staff, and especially their heads, Dr Isabelle Pellegrin, and Julien Jeanpetit (Centre Hospitalier Universitaire Robert Pellegrin, Bordeaux, France), Pr Edouard Tuaillon Centre de Ressources Biologiques du CHU de Montpellier), Dr Yves-Edouard Herpe (Centre de Ressources Biologiques Biobanque de Picardie), Pr Jacqueline Deloumeaux (Centre biologique du CHU de la Guadeloupe), Dr Rémi Neviere (CeRBiM, Centre de Ressources Biologiques de la Martinique), Julien Eperonnier, Estelle Nobecourt (Centre de Ressources Biologiques de la Réunion) for the quality of DBS sample management of the EpiCov study. We thank the biobank team in Inserm SC10, particularly Sophie Circosta. We also thank the staff of the UVE virology department, for the high-quality management of such a large number of serological assays.

We thank the staff of DREES and INSEE, for their collaboration in the implementation of the study, methodological input, sample selection, and the complex development of weights to correct for non-response.

We thank the Ipsos staff, including Christophe David and Valérie Blineau in particular, for their major contribution to the quality of data collection.

