## Supplementary Material for "Risk factors for household transmission of SARS-Cov-2: a modelling study in the French national population-based EpiCov cohort"

### Table S1. Size of the household in which individuals lived according to age

|  | **Age, years** | | | | | | | | | | |
| --- | --- | --- | --- | --- | --- | --- | --- | --- | --- | --- | --- |
|  | **06-10** | **11-14** | **15-17** | **18-24** | | **25-34** | **35-44** | **45-54** | **55-64** | **65-74** | **>74** |
|  | **N = 632** | **N = 832** | **N = 745** | **N = 1,797** | | **N = 1,304** | **N = 1,513** | **N = 3,100** | **N = 3,601** | **N = 3,244** | **N = 1,215** |
| **Household size** |  |  |  |  | |  |  |  |  |  |  |
| **1** | 0 (0) | 0 (0) | 0 (0) | 10.0 (180) | | 21.1 (275) | 13.2 (200) | 8.8 (274) | 13.1 (470) | 14.2 (462) | 21.0 (255) |
| **2** | 3.2 (20) | 2.9 (24) | 4.6 (34) | 14.5 (261) | | 51.5 (672) | 21.5 (325) | 23.7 (734) | 62.0 (2,233) | 79.2 (2,569) | 71.4 (868) |
| **3** | 17.1 (108) | 14.9 (124) | 18.7 (139) | 24.2 (435) | | 14.2 (185) | 15.9 (241) | 24.1 (748) | 16.6 (598) | 5.4 (174) | 5.8 (71) |
| **> 3** | 79.7 (504) | 82.2 (684) | 76.8 (572) | 51.3 (921) | | 13.2 (172) | 49.4 (747) | 43.4 (1,344) | 8.3 (300) | 1.2 (39) | 1.7 (21) |
| **Family structure** |  |  |  |  |  | |  |  |  |  |  |
| **Living alone** | 0 (0) | 0 (0) | 0 (0) | 10.0 (180) | 21.1 (275) | | 13.2 (200) | 8.8 (274) | 13.1 (470) | 14.2 (462) | 21.0 (255) |
| **Couple without children** | 0 (0) | 0 (0) | 0 (0) | 7.2 (129) | 46.9 (611) | | 17.8 (269) | 19.4 ( 601) | 58.9 (2,120) | 76.8 (2,492) | 68.0 (826) |
| **Single-parent family** | 9.0(57) | 11.4 (95) | 11.7 (87) | 11.5 (206) | 4.2 (55) | | 5.6 (84) | 6.2 ( 192) | 2.7 (99) | 1.0 ( 3.1) | 2.6 (32) |
| **Couple with one or more children** | 89.7(567) | 86.8 (722) | 86.0 (641) | 65.3(1,173) | 21.2 (277) | | 60.9 (922) | 63.1 (1,956) | 22.9 (826) | 5.1 (164) | 3.8 (46) |
| **3 generation family** | 0.8 (5) | 1.2 (10) | 1.1 (8) | 0.6 (10) | 0.7 (9) | | 0.5 (8) | 0.5 (17) | 0.2 (9) | 0.3 (9) | 1.2 (14) |
| **Other household structure** | 0.5 (3) | 0.6 (5) | 1.2 (9) | 5.5 (99) | 5.9 (77) | | 2.0 (30) | 1.9 (60) | 2.1 (77) | 2.7 (86) | 3.5 (42) |
| Results are shown as % (n) | | | | | | | | | | | |

### Table S2. Number of recruited and seropositive individuals by age and household size

| **variable** | **Overall  n / N  (95%CI)** | **Size 1  n / N  (95%CI)** | **Size 2  n / N  (95%CI)** | **Size 3  n / N  (95%CI)** | **Size 4  n / N  (95%CI)** | **Size >4  n / N  (95%CI)** |
| --- | --- | --- | --- | --- | --- | --- |
| **HOUSEHOLDS** | N = 8,165 | N = 2,116 | N = 3,870 | N = 941 | N = 937 | N = 301 |
| **≥ 1 seropositive** | 784 | 120 | 302 | 124 | 172 | 66 |
|  | 9.6 (9.0 - 10.0) | 5.7 (4.7 - 6.8) | 7.8 (7.0 - 8.7) | 13.0 (11.0 - 16.0) | 18.0 (16.0 – 21.0) | 22.0 (17.0 – 27.0) |
| **> 1 seropositive** | 229 | 0 | 95 | 41 | 59 | 34 |
|  | 2.8 (2.5 – 3.2) | - | 2.5 (2.0 – 3.0) | 4.4 (3.2 - 5.9) | 6.3 (4.9 -8.1) | 11.0 (8.1 – 16.0) |
| **INVIDIVUALS** | N = 17,983 | N = 2,116 | N = 7,740 | N = 2,823 | N = 3,748 | N = 1,556 |
| **Overall** | 1,107 | 120 | 397 | 175 | 277 | 138 |
|  | 6.2 (5.8 – 6.5) | 5.1 (4.7- 6.8) | 5.1 (4.7 - 5.7) | 6.2 (5.4 – 7.2) | 7.4 (6.6 - 8.3) | 8.9 (7.5 – 10.0) |
| **Age, years** |  |  |  |  |  |  |
| **06-10** | 35 / 632 | 0 / 0 | 1 / 20 | 5 / 108 | 19 / 337 | 10 / 167 |
|  | 5.5 (3.9 - 7.7) | - | 5.0 (0.3 – 27.0) | 4.6 (1.7 – 11.0) | 5.6 (3.5 – 8.8) | 6.0 (3.1 – 11.0) |
| **11-14** | 62 / 832 | 0 / 0 | 2 / 24 | 12 / 124 | 27 / 452 | 21 / 232 |
|  | 7.5 (5.8 – 9.5) | - | 8.3 (1.5 – 28.0) | 9.7 (5.3 – 17.0) | 6.0 (4.0 – 8.7) | 9.1 (5.8 – 14.0) |
| **15-17** | 57 / 745 | 0 / 0 | 5 / 34 | 7 / 139 | 26 / 373 | 19 / 199 |
|  | 7.7 (5.9 – 9.9) | - | 15.0 (5.5 - 32.0) | 5.0 (2.2 – 10.0) | 7.0 (4.7 – 10.0) | 9.5 (6.0 – 15.0) |
| **18-24** | 192 / 1,797 | 21 / 180 | 22 / 261 | 45 / 435 | 66 / 615 | 38 / 306 |
|  | 11.0 (9.3 - 12.0) | 12.0 (7.5 – 17.0) | 8.4 (5.5 - 13.0) | 10.0 (7.7 – 14.0) | 11.0 (8.5 – 14.0) | 12.0 (9.0 – 17.0) |
| **25-34** | 101 / 1,304 | 23 / 275 | 55 / 672 | 11 / 185 | 8 / 122 | 4 / 50 |
|  | 7.7 (6.4 – 9.4) | 8.4 (5.5 – 12.0) | 8.2 (6.3 – 11.0) | 5.9 (3.2 – 11.0) | 6.6 (3.1 – 13.0) | 8.0 (2.6 – 20.0) |
| **35-44** | 95 / 1,513 | 9 / 200 | 19 / 325 | 11 / 241 | 45 / 570 | 11 / 177 |
|  | 6.3 (5.1 - 7.7) | 4.5 (2.2 – 8.6) | 5.8 (3.7 – 9.1) | 4.6 (2.4 – 8.2) | 7.9 (5.9 – 10.0) | 6.2 (3.3 – 11.0) |
| **45-54** | 205 / 3,100 | 18 / 274 | 43 / 734 | 43 / 748 | 69 / 1,000 | 32 / 344 |
|  | 6.6 (5.8 – 7.6) | 6.6 (4.1 – 10.0) | 5.9 (4.3 – 7.9) | 5.7 (4.2 – 7.7) | 6.9 (5.4 – 8.7) | 9.3 (6.5 – 13.0) |
| **55-64** | 180 / 3,601 | 22 / 470 | 111 / 2,233 | 30 / 598 | 14 / 237 | 3 / 63 |
|  | 5.0 (4.3 - 5.8) | 4.7 (3.0 – 7.1) | 5.0 (4.1 – 6.0) | 5.0 (3.5 – 7.2) | 5.9 (3.4 – 9.9) | 4.8 (1.2 – 14.0) |
| **65-74** | 143 / 3,244 | 16 / 462 | 117 / 2,569 | 8 / 174 | 2 / 30 | 0 / 9 |
|  | 4.4 (3.7 – 5.2) | 3.5 (2.1 – 5.7) | 4.6 (3.8 – 5.5) | 4.6 (2.2 – 9.2) | 6.7 (1.2 – 24.0) | - |
| **>74** | 37 / 1,215 | 11 / 255 | 22 / 868 | 3 / 71 | 1 / 12 | 0 / 9 |
|  | 3.0 (2.2 – 4.2) | 4.3 (2.3 – 7.8) | 2.5 (1.6 – 3.9) | 4.2 (1.1 – 13.0) | 8.3 (0.4 – 40.0) | - |
| 95% CI, 95% confidence interval ^a^ 16 missing values for gender | | | | | | |

### Table S3. Comparison of model performance and estimated parameters

Factors that may be associated with the risk of infection from extra-household exposure and/or a single infected household member were progressively added to the model.

Lower WAIC and LOOIC scores indicate better model fit.

| **A) Characteristics of the susceptible individual** | | | | |
| --- | --- | --- | --- | --- |
| **Variable** | **Model 1** | | **Model 2** | **Model 3** |
|  | **extra-household** | **intra-household** | **extra-household** | **intra-household** |
| **Age of the susceptible, years** |  |  |  |  |
| **06-10** | 0.9 (0.5 - 1.5) | 0.4 (0.2 - 0.8) |  |  |
| **11-14** | 1.2 (0.8 - 1.8) | 0.7 (0.4 - 1.3) |  |  |
| **15-17** | 1.5 (1.0 - 2.2) | 0.7 (0.3 - 1.4) |  |  |
| **18-24** | 2.5 (2.0 - 3.3) | 0.5 (0.3 - 0.9) |  |  |
| **25-34** | 1.9 (1.4 - 2.5) | 0.5 (0.2 - 1.0) |  |  |
| **35-44** | 1.3 (1.0 - 1.8) | 0.5 (0.2 - 0.9) |  |  |
| **45-54** | 1.1 (0.8 - 1.4) | 0.8 (0.5 - 1.3) |  |  |
| **55-64** | ref | ref |  |  |
| **65-74** | 0.8 (0.6 - 1.1) | 2.7 (1.5 - 4.7) |  |  |
| **>74** | 0.7 (0.5 - 1.0) | 0.8 (0.2 - 2.0) |  |  |
| **Sex of the susceptible** |  |  |  |  |
| **Female** |  |  | - | - |
| **Male** |  |  | 0.9 (0.8 - 1.1) | 0.7 (0.5 - 1.0) |
| **N** | 7,291.2 (19.8) | | 17,950 | 17,950 |
| **WAIC (p_waic)** | -52.5 | | 7,379.5 (3) | 7,378.2 (2.9) |
| **Delta WAIC / model null** | 7,291.3 (19.9) | | 0.1 | -0.6 |
| **LOOIC (p_loo)** | -52.4 | | 7,379.5 (3) | 7,378.2 (2.9) |
| **Delta LOO-PSIS / model null** | 7,291.2 (19.8) | | 0.1 | -0.6 |

| **B) Characteristics of the potential infector** | | | |
| --- | --- | --- | --- |
| **Variable** | **Model 4** | **Model 5** | **Model 6** |
|  | **intra-household** | **intra-household** | **intra-household** |
| **Age of the infector, years** |  |  |  |
| **06-10** | 0.5 (0.2 - 1.1) |  |  |
| **11-14** | 0.5 (0.2 - 1.1) |  |  |
| **15-17** | 0.11 (0.02 - 0.34) |  |  |
| **18-24** | 0.15 (0.05 - 0.31) |  |  |
| **25-34** | 0.3 (0.1 - 0.7) |  |  |
| **35-44** | 0.3 (0.1 - 0.6) |  |  |
| **45-54** | 0.9 (0.5 - 1.4) |  |  |
| **55-64** | ref |  |  |
| **65-74** | 1.5 (0.8 - 2.6) |  |  |
| **>74** | 0.7 (0.2 - 2.0) |  |  |
| **Sex of the infector** |  |  |  |
| **Female** |  | ref |  |
| **Male** |  | 0.8 (0.5 - 1.3) |  |
| **Family ties** |  |  |  |
| **Between partners** |  |  | ref |
| **From mother to child** |  |  | 1.0 (0.6 - 1.5) |
| **From father to child** |  |  | 0.4 (0.1 - 0.7) |
| **From child < 12 years old to parent** |  |  | 0.31 (0.06 - 0.81) |
| **From child ≥ 12 years old to parent** |  |  | 0.10 (0.02 - 0.24) |
| **From grandparent to grandchild** |  |  | 0.53 (0.04 - 4.78) |
| **From grandchild to grandparent** |  |  | 0.39 (0.04 - 2.59) |
| **From a sibling < 12 years old** |  |  | 0.39 (0.07 - 1.14) |
| **From a sibling ≥ 12 years old** |  |  | 0.3 (0.2 - 0.6) |
| **Between individuals with other family ties** |  |  | 0.4 (0.1 - 1.3) |
| **Between individuals with no family ties** |  |  | 0.13 (0.02 - 0.66) |
| **N** | 17,983 | 17,950 | 17,983 |
| **WAIC (p_waic)** | 7,337.5 (10.8) | 7,380.9 (3.4) | 7,339.9 (8.8) |
| **Delta WAIC / model null** | -29.3 | 0.8 | -28.1 |
| **LOOIC (p_loo)** | 7,337.5 (10.8) | 7,380.9 (3.4) | 7,340 (8.9) |
| **Delta LOO-PSIS / model null** | -29.3 | 0.8 | -28.1 |

| **C) Household characteristics** | | | | |
| --- | --- | --- | --- | --- |
| **Variable** | **Model 7** | **Model 8** | **Model 9** | **Model 10** |
|  | **extra-household** | **intra-household** | **extra-household** | **intra-household** |
| **Family income** |  |  |  |  |
| **D01 (lowest)** | 1.2 (0.8 - 1.8) | 0.5 (0.2 - 1.2) |  |  |
| **D02-D03** | 1.0 (0.7 - 1.4) | 1.3 (0.7 - 2.3) |  |  |
| **D04-D05** | ref | ref |  |  |
| **D06-D07** | 1.2 (0.9 - 1.5) | 1.1 (0.7 - 1.9) |  |  |
| **D08-D09** | 1.3 (1.1 - 1.7) | 1.1 (0.7 - 1.7) |  |  |
| **D10 (highest)** | 1.7 (1.3 - 2.2) | 1.4 (0.9 - 2.3) |  |  |
| **Population density in the municipality of residence** |  |  |  |  |
| **Low** |  |  | ref | ref |
| **Medium** |  |  | 1.2 (1.0 - 1.4) | 1.0 (0.7 - 1.4) |
| **High** |  |  | 1.6 (1.4 - 1.9) | 1.0 (0.7 - 1.4) |
| **N** | 17,704 | 17,704 | 17,983 | 17,983 |
| **WAIC (p_waic)** | 7,261.7 (7) | 7,277.2 (7.4) | 7,366.3 (4.1) | 7,400.4 (4.4) |
| **Delta WAIC / model null** | -6.3 | 1.5 | -14.9 | 2.1 |
| **LOOIC (p_loo)** | 7,261.7 (7) | 7,277.2 (7.4) | 7,366.3 (4.1) | 7,400.4 (4.4) |
| **Delta LOO-PSIS / model null** | -6.3 | 1.5 | -14.9 | 2.1 |
| **Variable** | **Model 11** | **Model 12** |  |  |
|  | **extra-household** | **extra-household** |  |  |
| **Living in a socially deprived neighbourhood** |  |  |  |  |
| **No** | ref |  |  |  |
| **Yes** | 1.0 (0.6 - 1.7) |  |  |  |
| **Region** |  |  |  |  |
| **Auvergne-Rhone-Alpes** |  | 1.9 (1.3 - 2.9) |  |  |
| **Bourgogne-Franche-Comte** |  | 1.1 (0.7 - 1.9) |  |  |
| **Bretagne** |  | 0.6 (0.3 - 1.0) |  |  |
| **Centre-Val de Loire** |  | ref |  |  |
| **Corse** |  | 0.24 (0.02 - 1.22) |  |  |
| **Grand Est** |  | 1.5 (1.0 - 2.3) |  |  |
| **Hauts-de-France** |  | 1.8 (1.2 - 2.7) |  |  |
| **Ile-de-France** |  | 2.3 (1.6 - 3.5) |  |  |
| **Normandie** |  | 1.1 (0.7 - 1.9) |  |  |
| **Nouvelle-Aquitaine** |  | 0.8 (0.5 - 1.3) |  |  |
| **Occitanie** |  | 0.9 (0.6 - 1.4) |  |  |
| **Pays de la Loire** |  | 1.0 (0.6 - 1.6) |  |  |
| **Provence-Alpes-Cote d'Azur** |  | 0.8 (0.5 - 1.4) |  |  |
| **N** | 17,983 | 17,983 |  |  |
| **WAIC (p_waic)** | 7,398 (3.1) | 7,287.2 (12.9) |  |  |
| **Delta WAIC / model null** | 0.9 | -54.5 |  |  |
| **LOOIC (p_loo)** | 7,398 (3.1) | 7,287.2 (12.9) |  |  |
| **Delta LOO-PSIS / model null** | 0.9 | -54.5 |  |  |

| **Variable** | **Model 13** | **Model 14** | **Model 15** |
| --- | --- | --- | --- |
|  | **intra-household** | **intra-household** | **intra-household** |
| **Accommodation type** |  |  |  |
| **A house with a yard or a garden** | ref |  |  |
| **A house with no yard or garden** | 1.5 (0.4 - 4.0) |  |  |
| **An apartment with a balcony or a terrace** | 1.3 (0.9 - 1.8) |  |  |
| **An apartment with a community garden** | 0.8 (0.1 - 2.5) |  |  |
| **An apartment with no balcony, terrace or community garden** | 1.1 (0.5 - 2.1) |  |  |
| **Other** | 0.9 (0.2 - 3.1) |  |  |
| **Household size** |  |  |  |
| **2** |  | ref |  |
| **3** |  | 0.5 (0.3 - 0.7) |  |
| **4** |  | 0.4 (0.3 - 0.6) |  |
| **>4** |  | 0.4 (0.3 - 0.6) |  |
| **Overcrowded housing, defined as at least two people living in less than 18 m^2^ per person** |  |  |  |
| **Crowded housing** |  |  | 1.2 (0.8 - 1.7) |
| **Housing not particularly crowded** |  |  | ref |
| **N** | 17,980 | 17,983 | 16,883 |
| **WAIC (p_waic)** | 7,403 (7) | 7,373.3 (5.4) | 6,739.2 (3.3) |
| **Delta WAIC / model null** | 3.6 | -11.4 | 0.7 |
| **LOOIC (p_loo)** | 7,403.3 (7.1) | 7,373.3 (5.5) | 6,739.2 (3.3) |
| **Delta LOO-PSIS / model null** | 3.7 | -11.4 | 0.7 |
| **Variable** | **Model 17** | **Model 18** |  |
|  | **extra-household** | **intra-household** |  |
| **Immigration history of the respondent** |  |  |  |
| **Majority population** | ref | ref |  |
| **1st-generation immigrant from Europe** | 0.8 (0.5 - 1.2) | 1.0 (0.4 - 2.1) |  |
| **1st-generation immigrant from outside Europe** | 1.6 (1.0 - 2.4) | 0.6 (0.2 - 1.2) |  |
| **2nd-generation immigrant from Europe** | 1.0 (0.7 - 1.4) | 1.3 (0.7 - 2.3) |  |
| **2nd-generation immigrant from outside Europe** | 1.6 (1.0 - 2.3) | 1.3 (0.7 - 2.4) |  |
| **N** | 17,442 | 17,442 |  |
| **WAIC (p_waic)** | 7,187.2 (6.1) | 7,196 (7.6) |  |
| **Delta WAIC / model null** | -1.3 | 3.0 |  |
| **LOOIC (p_loo)** | 7,187.2 (6.1) | 7,196.2 (7.7) |  |
| **Delta LOO-PSIS / model null** | -1.3 | 3.1 |  |
| Median 95% Crl: 95% Credible interval | | |  |
| Abbreviations: LOOIC, LOO information criterion; WAIC, Watanabe-Akaike information criterion; p_loo and p_waic, effective number of parameters for estimation of LOOIC and WAIC, respectively | | | |

### Supplementary Note 1: The COVID-19 pandemic in France in 2020

The first wave of the COVID-19 pandemic peaked two weeks after the first national lockdown decreed from March 17 to May 11, in the context of mask shortages and little availability of PCR tests. This first lockdown combined drastic measures, including limited outdoor circulation, travel bans, mandatory teleworking, and the closure of schools, universities, and shops, except for essential supplies, which led to a very low incidence rate. The second wave started slowly at the end of August, despite the wide-scale distribution of masks and free access to PCR and antigenic tests. Following a curfew period with territorial variations, a second national lockdown was reinstated from October 30 to December 15, 2020. It was less restrictive than the previous lockdown, with no school closures (although universities were closed) and an extended list of shops were authorized to remain open. Throughout the year, incentives for telework and other barrier measures, especially face covering and physical distancing, were maintained.

### Supplementary Note 2: Technical summary

We adopted an adapted version of chain-binomial models that fit the final size of infections^1^, i.e., the distribution of seropositive and seronegative individuals within households, to analyze the transmission process among household members.

The model has been previously described by Bi et al. (2020)^2^. We adapted their code available in open access to the EpiCov data.

The model estimates the risk of infection from: 1) extra-household sources and 2) a single infected household member.

#### Assumptions

The model’s assumptions are as follows:

- each household member can be infected either from within a household or from extra-household sources

- household members mix at random within a household and can infect one another

- all household members were initially susceptible to infection to SARS-CoV-2

- the possibility of reinfection for the duration of the study period was neglected

In addition, we assumed no misclassification of the serological result, either positive or negative.

#### Data augmentation

Given that only the serological status of individuals was known, with no additional information about the chronology of infection events within the households, the model considers augmented data with all possible sequences of viral introductions to each household and subsequent transmission events within the household.

Possible sequences of viral introduction in the household and subsequent transmission events within the household are defined from the assignment of a generation to each household member. People infected from outside the household are assigned to generation 0. Those that they infect within the household are assigned to generation 1, those infected by generation 1 to generation 2, and so on. Uninfected individuals are assigned to generation infinity. For each household *h*, one *k* possible sequence ${HH}_{h,k}$ of viral introduction/transmission is one ordered assignment of generation.

For example, in a household of 3 individuals, *i, j,* and *k*, in which the 2 individuals *i* and *k* are positive and *j* is negative, there are three possible sequences (Fig S2).

In the first, ${HH}_{1}$, both *i* and *k* could have been infected outside of the household: the two are assigned to generation 0.

In the second, ${HH}_{2}$, *i* could have been infected outside and then infected *k* within the household: *i* is assigned to generation 0 and *k* to generation 1.

The third, ${HH}_{3}$, is the opposite: *k* could have been infected outside and then infected *i* within the household: *k* is assigned to generation 0 and *i* to generation 1.

In these three sequences, *j* is assigned to generation infinity.

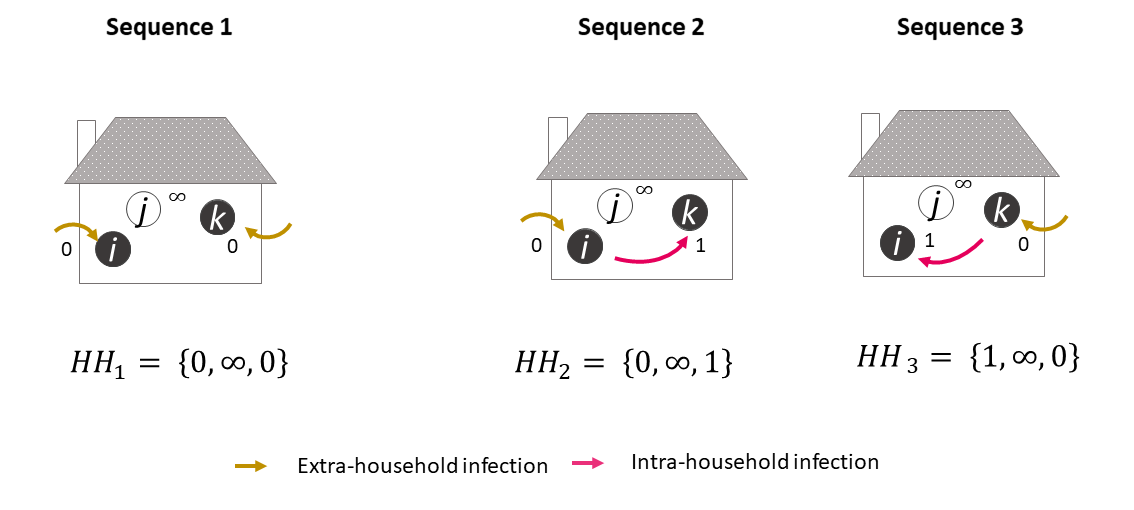

**Figure S1. All possible sequences of viral introduction to each household and subsequent transmission events within the household for a household with three members with two positive individuals.**

#### Likelihood of the model

The likelihood of the model is calculated via the decomposition into the contribution of each possible sequence ${HH}_{h,k}$ of each household *h*.

The likelihood of the sequence ${HH}_{h,k}$ is: $\Pr\left( {HH}_{h,k} \right)=\prod_{i} Pr(g_{i}|{HH}_{h,k})$

where $\Pr\left( g_{i} | \mathrm{HH}_{h,k} \right)$ is the probability of household member *i* of household *h* having an infection generation of $g_{i}$ in the sequence $\mathrm{HH}_{h,k}$

$\Pr\left( g_{i} | \mathrm{HH}_{h,k} \right)$ is defined from the two probabilities of interest:

1) The probability $Q_{i,j}$ of a household member *i* escaping infection from a single infectious household member *j*, which corresponds to a person-to-person transmission probability.

2) The probability $B_{i}$ of a household member *i* escaping infection from the community, i.e., extra-household exposure, over the course of epidemic.

$$\Pr\left( g_{i} | \mathrm{HH}_{h,k} \right)=\left( 1-B_{i} \right)^{I\left( g_{i}=0 \right)}\left( B_{i} \right)^{I\left( g_{i}\neq0 \right)}[\prod_{j\neq i,g_{j}<\left( g_{i}-1 \right)} Q_{i,j}][1-\prod_{j\neq i,g_{j}=(g_{i}-1)} Q_{i,j}]$$

with $I:=\left\{ \begin{aligned} 1, g_{i}=0 \\ 0, g_{i}\neq0 \end{aligned} \right.$

If i is infected outside, i.e., assigned to generation 0, it is simplified as $\Pr\left( g_{i} | \mathrm{HH}_{h,k} \right)=1-B_{i}$

If i is infected within the household, i.e., assigned to generation 1 or greater, it is simplified as

$$\Pr\left( g_{i} | \mathrm{HH}_{h,k} \right)=B_{i}[\prod_{j\neq i,g_{j}<\left( g_{i}-1 \right)} Q_{i,j}][1-\prod_{j\neq i,g_{j}=(g_{i}-1)} Q_{i,j}]$$

Where:

$B_{i}$ represents the probability of household member *i* escaping infection from extra-household exposure

$\prod_{j\neq i,g_{j}<\left( g_{i}-1 \right)} Q_{i,j}$ represents the probability of household member *i* escaping infection from other infected household members up to generation $g_{i}$

$1-\prod_{j\neq i,g_{j}=(g_{i}-1)} Q_{i,j}$ represents the probability of household member i being infected from any infected household members of generation $g_{i}-1$

For each household *h*, the likelihood of observing the final infection state is the sum of the probability of all the possible sequences ${HH}_{h,k}$ that could lead to this final result.

$$Pr({HH}_{h})=\sum_{k} Pr({HH}_{h,k})$$

The global log-likelihood of the model is the sum of the contribution of all households:

$$logLik=\sum_{h} log(\Pr\left( HH_{h} \right))$$

#### Covariates of adjustment

In a null model, the probabilities Q and B were fixed and equal for all individuals. Then, they were adjusted for individual and household characteristics.

$Q_{i,j}$ was estimated as a function of the exposed household member individual’s characteristics $\boldsymbol{X}_{\boldsymbol{i}}$, the potential infectors’ characteristics $\boldsymbol{X}_{\boldsymbol{j}}$, and some shared characteristics of their household $\boldsymbol{X}_{\boldsymbol{h}}$ as follows:

$$logit\left( Q_{i,j} \right)=\beta_{0}+\boldsymbol{X}_{\boldsymbol{i}}\boldsymbol{\beta}+\boldsymbol{X}_{\boldsymbol{j}}\boldsymbol{\alpha+}\boldsymbol{X}_{\boldsymbol{h}}\boldsymbol{\gamma}$$

$B_{i}$, was estimated as a function of the exposed household member individual’s characteristics $\boldsymbol{X}_{\boldsymbol{i}}$ and his household’s characteristics $\boldsymbol{X}_{\boldsymbol{h}}$**.**

$$logit\left( B_{i} \right)=\beta_{0}'+\boldsymbol{X}_{\boldsymbol{i}}\boldsymbol{\beta'+}\boldsymbol{X}_{\boldsymbol{h}}\boldsymbol{\gamma}\mathbf{'}$$

The probabilities of being infected from the community and from one single infected household member were then obtained as $1-expit(logit\left( B_{i} \right)$) and $1-expit(logit\left( Q_{i,j} \right)$) respectively, the expit function being the inverse of logit.

We consider the following covariates.

- **Covariates affecting susceptibility,** $\boldsymbol{X}_{\boldsymbol{i}}$

Characteristics of the exposed (or susceptible) individual i

- age group (categorical): 6-10, 11-14, 15-17, 18-24, 25-34, 35-44, 45-54, 55-64, 65-74, >74

- gender (binary): female/male

- **Covariates affecting Infectivity,** $\boldsymbol{X}_{\boldsymbol{j}}$

Characteristics of the potential infector j

- age group (categorical): 6-10, 11-14, 15-17, 18-24, 25-34, 35-44, 45-54, 55-64, 65-74, >74

- gender (binary): female/male

- family relationship with the individual *i* (categorical): partner/spouse, mother, father, child < 12 years old, child ≥ 12 years old, grandparent, grandchild < 12 years old, grandchild ≥ 12 years old, sibling < 12 years old, sibling ≥ 12 years old, other family link, no family link

- **Household-level covariates,** $\boldsymbol{X}_{\boldsymbol{k}}$

- family income in deciles (categorical): D01 (lowest), D02-D03, D04-05, D06-07, D06-09, D10 (highest)

- density population in the municipality of residence (categorical): low, medium, high

- living in a socially deprived neighborhood (binary)

- accommodation type (categorical): an apartment with no balcony, terrace, or community garden, an apartment with a balcony or a terrace, an apartment with a community garden

house with a yard or a garden, a house with a yard or a garden, other

- overcrowded housing, defined as less than 18 m^2^ per inhabitant (binary)

- household size (categorical): 1, 2, 3, 4, ≥ 5 individuals

- region (categorical): the 13 administrative regions of mainland France

- immigration history (categorical): majority population, 1st-generation immigrant from Europe, 2nd-generation immigrant from Europe, 1st-generation immigrant from outside Europe, 2nd-generation immigrant from outside Europe. As the migration history was available only for the respondent member of the household and not all household members, this information was treated as a household-level covariate

#### Model selection

Associations of all the covariates mentioned with $B_{i}$ and $Q_{i,j}$, respectively, were tested one by one in univariate models. In the final multivariate model, we adjusted for covariates for which a decrease was observed in the widely applicable information criterion (WAIC) and the leave-one-out cross-validation information criterion (LOOIC) in univariate analyses ^21^.

#### Inference and implementation

Posterior distributions of parameters were estimated via MCMC using the rstan package. The default algorithm in rstan is the No-U-Turn Sampler (NUTS), which is a tuning-free Hamiltonian-based Monte Carlo sampler^3^.

We set weakly informative priors on all parameters to be normally distributed on the logit scale with a mean of 0 and a standard error of 1.5. We ran four chains of 1,500 iterations each, with 500 warm-up iterations, and assessed convergence visually and using the Gelman-Rubin Convergence Statistic (R-hat).

#### Handling of missing variables

Given the very low percentage of missing data for the considered variables (< 4%), models were run using the complete dataset.

#### Simulation of source of infection

For each household with at least one seropositive individual, we drew one sequence of viral introduction and subsequent within-household transmission from the probability distribution of all possible sequences of the household. We then estimated the number of infections acquired from extra-household exposure and the number of within-household transmission events in the drawn scenario.
